# County-Specific, Real-Time Projection of the Effect of Business Closures on the COVID-19 Pandemic

**DOI:** 10.1101/2021.02.10.21251533

**Authors:** Dominic Yurk, Yaser Abu-Mostafa

**Affiliations:** Department of Electrical Engineering, California Institute of Technology, Pasadena, CA 91125

**Keywords:** data-driven, model, COVID-19, policy, neural-net

## Abstract

Public health policies such as business closures have been one of our most effective tools in slowing the spread of COVID-19, but they also impose costs. This has created demand from policy makers for models which can predict when and where such policies will be most effective to head off a surge and where they could safely be loosened. No current model combines data-driven, real-time policy effect predictions with county-level granularity. We present a neural net-based model for predicting the effect of business closures or re-openings on the COVID-19 time-varying reproduction number *R*_*t*_ in real time for every county in California. When trained on data from May through September the model accurately captured relative county dynamics during the October/November California COVID-19 surge (*r*^2^ = 0.76), indicating robust out-of-sample performance. To showcase the model’s potential utility we present a case study of various counties in mid-October. Even when counties imposed similar restrictions at the time, our model successfully distinguished counties in need of drastic and immediate action to head off a surge from counties in less dire need of intervention. While this study focuses on business closures in California, the presented model architecture could be applied to other policies around world.

## 1 Introduction

From the beginning of the COVID-19 pandemic it was recognized that public health policy would be our most effective tool in limiting the spread of the disease. However, while some measures such as social distancing were quickly and widely agreed upon, other policies such as business closures have proven far more contentious. While the economic impact of business closures is immediately evident, the corresponding amount of public health benefit cannot be directly calculated. In order to make informed decisions policy makers have turned to model-based analyses which attempt to estimate how effective various policies are at slowing the spread of COVID-19. Early in the pandemic, the lack of information on the spread of this new disease made truly data-driven analyses of policy effectiveness impossible. Attempts to model policy impacts were forced to rely on assumptions to make up for the scarcity of available data, using techniques such as curve fitting and SEIR models that impose constraints on disease evolution [1–6]. Unfortunately the pandemic has continued to spread widely over the past year, leading to multiple rounds of tightening, loosening, and re-tightening of public health polices around the world. The variation in timing and austerity of these policies between and within countries has created a variety of natural experiments, allowing researchers to model policy effectiveness in a more data-driven way.

To date, most data-driven policy analyses have been retrospective and focused on overall effectiveness across a number of counties, states, or countries [7–10]. These analyses can be broadly useful for policy makers deciding which controls to apply when attempting to contain an outbreak already in progress. However, such models are unable to identify how policy effects differ in different areas, overlooking the fact that optimal policy may vary greatly between urban centers and rural communities or between an area with one daily case per 10,000 residents and an area with 100. Furthermore, due to long delays between initial infection and subsequent transmission, testing, and reporting, by the time a COVID-19 outbreak becomes readily apparent even an immediate and strict lockdown will take substantial time to contain it. For example, in the United States the strictest lockdown imposed to date was in New York City on March 22nd, but average daily cases did not peak until almost 3 weeks later on April 11th [11]. Thus, there is a need for a real-time, locality-specific model of policy effectiveness capable of alerting officials to the need for action before an outbreak is well underway.

An attempt to fill this gap has recently been presented which projects future mortality rates in various countries under different policy scenarios using Gaussian processes built on top of an SEIR model [12]. However, the only policy variable used is a “stringency index” which clusters many policies from business and school closures to mask wearing and international travel restrictions across an entire country [13]. This not only obscures the effect of individual policies, but would also be of limited use in a country like the United States where policy varies widely between localities. More detailed analysis has largely been frustrated by the lack of datasets capturing county-level public health policies. This problem has recently been addressed by the Stanford COVID-19 simulation group (SC-COSMO), which released a detailed dataset covering county-level public health policies across California [14]. We present a new model based on this data which uses monotonic neural networks to perform real-time county-level policy effect forecasting in California. In this study we focus on the effect of business closures and present both a test data set supporting model accuracy and a case study demonstrating potential utility to policy makers. The structure of our model is simple and relies on only a few input parameters, making it potentially applicable to other policies and other localities across the US and the world.

## 2 Methods

### 2.1 Input and Output Parameters

#### 2.1.1 Measuring *R*_*t*_

The model presented here focuses on the time-varying reproduction number *R*_*t*_ as the metric for assessing the state of the COVID-19 pandemic. *R*_*t*_ is a measure of how many new people an infected individual will transmit the virus to given that the individual was infected on day *t*. The primary advantage of this metric is that it is directly impacted by policy changes in real time, compared to other metrics such as COVID-19 cases and deaths which can take weeks to respond to policy changes. Another advantage of *R*_*t*_ is that it is easily interpretable at a glance; any value above 1 indicates accelerating spread, while any value below 1 indicates decelerating spread. This makes it an ideal actionable parameter for policy makers. Thus, we chose to make the output target of our model on any given day the change in *R*_*t*_ from 7 days before that date to 7 days after, referred to hereafter as Δ *R*_*t*_ or del_Rt.

Because *R*_*t*_ cannot be measured directly, we used the Rt.live methodology to extract it from testing data [15]. While Rt.live only publishes *R*_*t*_ values at the state and national levels, its computations rely only on daily positive and negative testing volumes in each region, allowing for universal applicability provided appropriate data. We adapted their code to compute *R*_*t*_ values for each county in California based on county-level testing data gathered from COVID ActNow [16]. For the 15 out of 58 counties in which negative testing volumes were not reported on all relevant dates, the counties’ test positivity rates were assumed to match that of the state overall.

#### 2.1.2 Policy Data

The SC-COSMO public health dataset for California tracks seven different policy categories by county with an austerity score of 0 to 10 on each day, as listed in Table 1 (an eighth policy category, order_gath, was excluded because it is not tracked for many counties in the dataset). Each of these order types has a potential impact on the pandemic, and ideally our analysis would have incorporated many or all of them. However, to analyze the effect of a given policy in a data-driven way we needed many instances of the policy loosening and tightening across different counties and times. Only order_closure, which on average changed over 7 times per county, definitively met this criterion. Other orders with potentially enough data for analysis were order_shome and order_bubble. However, these policies rely on individual cooperation and are very difficult to enforce, resulting in inconsistent effectiveness. For example, in the two weeks surrounding the March stay-at-home order in Los Angeles median traffic to recreation and retail businesses fell from 92% of normal to 57% of normal, while the November stay-at-home order corresponded to a drop from 73% or normal to 67% of normal <u>[17].</u> For these reasons we chose to focus exclusively on order_closure for this analysis.

**Table 1:** Number of County-Level Policy Changes in the SC-COSMO Dataset (May 1st - Dec 1st)

| Order | Description | Total Changes | Tighter Changes | Looser Changes |
| --- | --- | --- | --- | --- |
| closure | Overall closure status | 408 | 169 | 239 |
| bubble | Social bubble restrictions | 134 | 106 | 28 |
| masks | Mask restrictions | 112 | 109 | 3 |
| shome | Stay at home restrictions | 84 | 52 | 32 |
| quarres | Resident quarantine restrictions | 32 | 21 | 11 |
| lab | Lab & tracing requirements | 30 | 23 | 7 |
| quarvis | Visitor quarantine restrictions | 2 | 1 | 1 |

#### 2.1.3 Model Inputs

Our model was constructed with 6 input variables: 1) population density, 2) number of new cases per 10,000 residents in the last 2 weeks, 3) *R*_*t*_ value 21 days ago, 4) difference between *R*_*t*_ values 21 and 28 days ago, 5) current value of order_closure, and 6) difference in order_closure between today and 7 days ago. Parameter 6 is intended to be the operative variable for policy makers, as it allows them to project the effects of potential policy decisions if they were made today. In parameters 3 and 4, a 3 week delay is inserted in order to simulate the “fog of war” present in real policy-making situations; on any given day the real-time Rt.live estimates have large error bars due to delays in transmission, testing, and reporting. As we look backwards these error bars become narrower due to increased data availability up to roughly 3 weeks in the past, at which point the uncertainty settles to a steady-state minimum. Thus, all parameters were chosen such that they would be available to users in real time with a high degree of confidence.

When constructing our model we considered data from May 1st, 2020 to December 1st, 2020. The May 1st cutoff was applied to exclude the early phases of the pandemic when testing was scarce, much of the general public was panicked, and many restrictive policies were enacted at once, all of which obscured individual policy effects. The Dec 1st cutoff was applied to provide a 4 week buffer prior to model development at the end of December, equivalent to the 3 week *R*_*t*_ input delay plus one week for determining the subsequent change in *R*_*t*_. Data points consisted of one snapshot per week of every county in California during this period (excluding those with fewer than 10 total cases in the previous week), for a total of 1387 data points. These data were divided into points before and after October 1st, and the latter points were held in reserve as an uncontaminated test set which was not used until after model optimization was performed (see results and discussion). This was done to ensure that we could obtain as clear of a picture as possible of model performance now and in the future when we are only able to train on data from the past.

### 2.2 Monotonic Neural Network

Over the past decade neural network models have achieved world-leading accuracy in tasks ranging from text, speech, and image recognition to predicting protein structure. This is due to the models’ ability to identify hidden patterns in data sets without researcher-imposed assumptions or constraints. In data-poor scenarios this lack of constraints can lead to poor extrapolation and occasionally nonsensical predictions when compared to SEIR models traditionally favored by epidemiologists. However, the COVID-19 pandemic has produced a body of data larger than any previously seen in epidemiology, allowing neural networks to attain high levels of performance in COVID-19 prediction first as supplements to SEIR models [18–20] and more recently as standalone models [21, 22]. These results encouraged us to pursue neural networks for the application presented here.

By machine learning standards, our data set of 1387 points is still relatively small, leading to a risk of overfitting. We mitigated this risk in two ways; first by limiting our inputs to the six parameters described above, and second by applying a monotonicity constraint to the output relative to change in order_closure. Monotonicity constraints have previously been shown to improve accuracy and robustness across a variety of machine learning models, particularly when trained on small data sets [23–26]. In our case the monotonicity assumption seems very applicable; it merely specifies that implementing a tighter policy today will never make Δ *R*_*t*_ worse than it would have been under a looser policy, but does not apply any constraints on the magnitude of the difference in Δ *R*_*t*_ between policies. We utilized an existing framework which allowed us to construct a standard dense neural network model with a monotonicity constraint applied to only one input [27]. We applied rectified linear unit activation between hidden layers and no activation on the output layer, removing any potential bias towards a sigmoidal output shape.

For model hyperparameter optimization we utilized data from May 1st through October 1st (943 total data points) and performed 5-fold cross-validation to estimate performance. We did not follow the standard practice of selecting a random subset of points for validation, as this would lead to very high correlations between some training and validation points; for example, a model trained on Orange County data from June 1st and 15th should do a very good job predicting Orange County’s behavior on June 8th. Instead, at each step we selected a random subset of 20% of all counties and used all data points from those counties for validation. The error function being minimized was RMS error between predicted and actual Δ *R*_*t*_, training was done with the Adam optimizer [28], and model parameters being optimized were number of hidden layers, hidden layer size, optimizer learning rate, optimizer weight decay factor, and number of training epochs.

## 3 Results

The parameters minimizing validation error were 3 hidden layers of 6 nodes each, a learning rate of 10^*−*3^, a weight decay factor of 10^*−*5^, and 20 training epochs. This yielded training and validation RMS errors of 0.0403 and 0.0445 respectively. Figure 1a shows an example result from one subset of validation counties, demonstrating that model predictions on training and validation data share similar distributions and similar correlations with true values.

**Figure 1:**
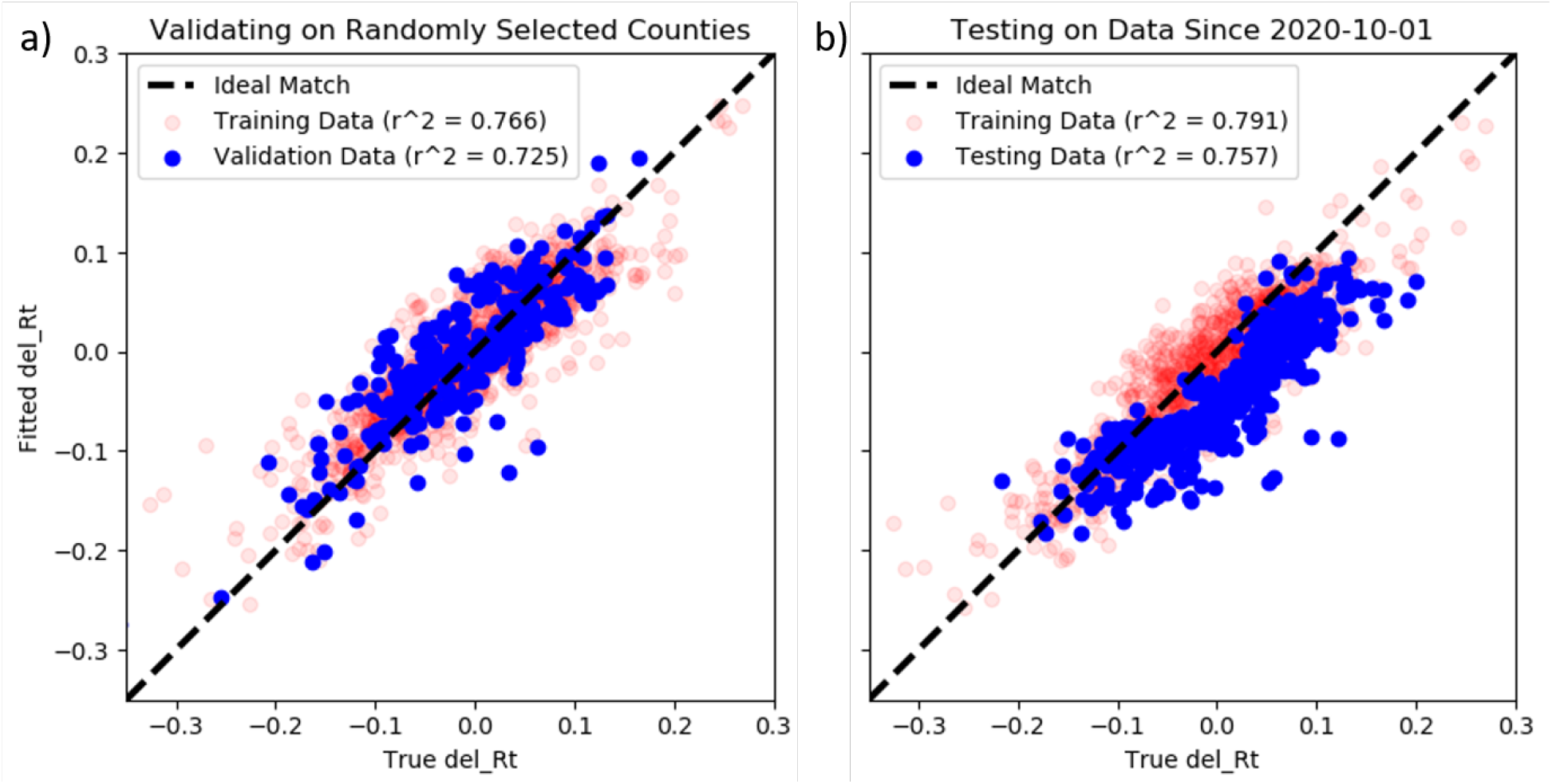
Model Validation and Testing. a) To optimize our model we validated on randomly withheld county data from May 1st through Oct 1st and chose model parameters to minimize validation error. b) We tested our model on data from Oct 2nd through Dec 1st. The resulting model outputs have a slight underprediction bias but still capture the overall trend of which counties have relatively good or bad *R*_*t*_ trends.

After completing hyperparameter optimization we trained our model on the full data set from May 1st to October 1st and tested its performance on our reserved data from October 2nd through December 1st (444 data points). This resulted in training and testing errors of 0.0411 and 0.0625 respectively. At first glance, these error values suggest that our model failed to generalize well to newer data. However, a closer look at all training and testing points in Figure 1b shows that correlation between predicted and true values is almost as good for the test set (*r*^2^ = 0.757) as it is for the training set (*r*^2^ = 0.791). The problem is that our model is biased towards underpredicting Δ *R*_*t*_ in the test set, particularly when the true Δ *R*_*t*_ is positive. This mirrors the fact that the ensemble of all major models tracked by the Reich Lab COVID-19 Forecasting Hub [29] consistently underpredicted future incident cases in California for epidemic weeks 41-50 (October 4th through December 6th), coinciding with by far the largest COVID-19 surge in California to date. The reasons for this surge are not yet well understood, but the consistent underprediction shows that it was driven by an unexpected factor that was not captured by almost any of the sophisticated models on the forecasting hub. Our model is relatively simple by design to avoid overfitting, so it should not be surprising that it is also unable to predict the full extent of this surge. However, the strong correlation between our model predictions and true values indicates that it could still provide valuable insights into the relative behavior of different regions under different policy options during such a period by performing county-specific policy impact predictions.

## 4 Discussion

The true utility of our model is its power to project the effects of a range of different policy options in real time for any county. Some randomly selected examples of these projections are shown in Figure 2. It is important to note that our model architecture requires these curves to be monotonically non-increasing, but it does not place any constraints on smoothness or shape. Thus, the observed smooth sigmoidal curves are an emergent property of the trained system. This matches our intuition for how the system should work; incremental policy changes produce incrementally smooth effects, and massive policy changes exhibit a law of diminishing returns. The diversity of centers and ranges in the sampled curves also demonstrates that the same policy change can have starkly different effects depending on when and where it is implemented.

**Figure 2:**
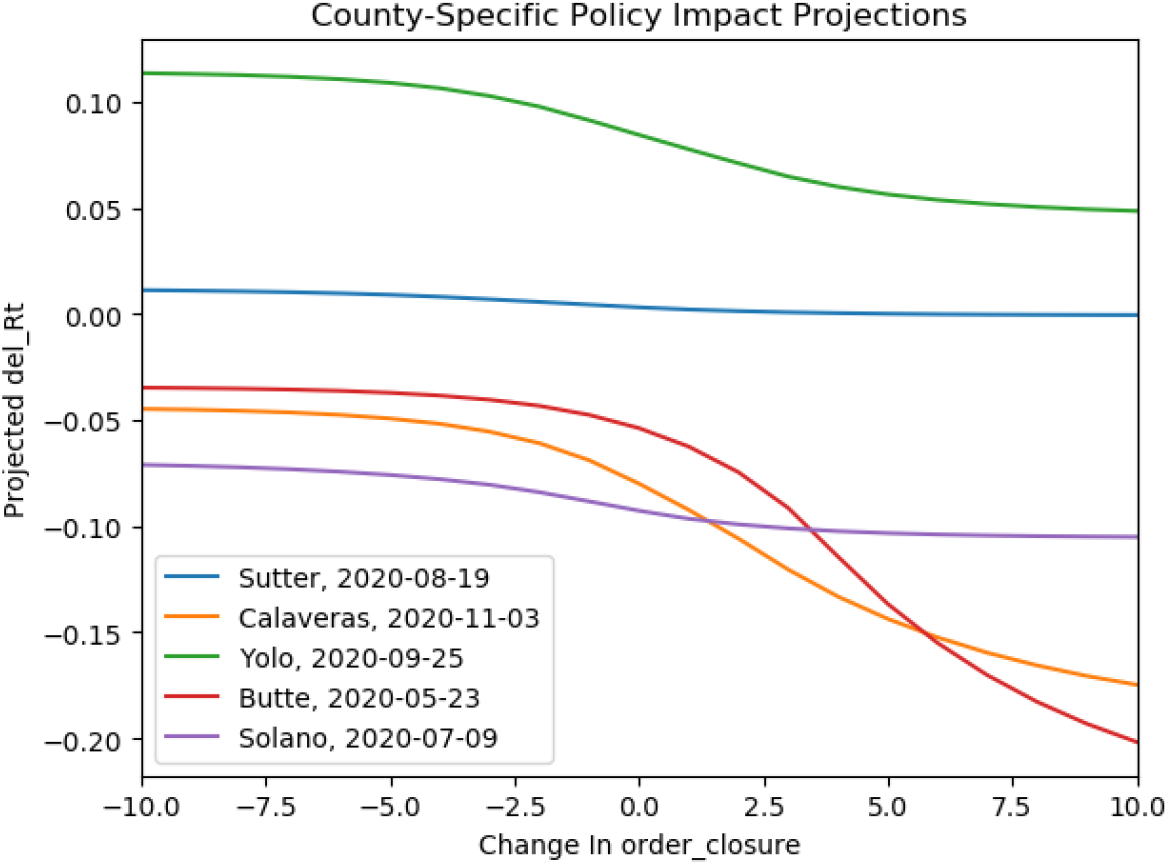
A Sample of County-Specific Policy Impact Predictions. We randomly selected five counties and times and show model projections of Δ *R*_*t*_ in each case under a range of possible policy changes. This shows that our model produces smooth sigmoid-shaped curves, in line with our intuition.

To demonstrate the potential power of this model we focus on the week of October 15th, which we now know was in the middle of a crucial period of rising *R*_*t*_ that later produced California’s devastating winter surge. At the time there was relatively little concern over COVID-19 trends; during this week eight counties loosened business closure restrictions and none tightened them. Using only data that were available at the time, our model would have strongly urged some of these counties to change course. We can broadly group the counties into three categories based on model projections; 1) those with a rapidly rising *R*_*t*_ trend which could be slowed but not reversed by business closures, 2) those with a rapidly declining *R*_*t*_ trend which would not reverse even with business reopenings, and 3) those at an inflection point where *R*_*t*_ could rise or fall depending on policy decisions. Figure 3 highlights one representative of each category in the form of Fresno, Mono, and Placer counties respectively. All three counties had relatively loose business closure policies for all of October; Fresno was at 3, Mono was at 2, and Placer dropped from 3 to 2 early in the month. Our model would have suggested that such policy was appropriate in Mono, which showed a substantial decrease in *R*_*t*_ after October 15th. In contrast, Fresno was at the opposite end of the spectrum during that same week with a rapidly rising *R*_*t*_. Our model would have projected that business closures would help slow this trend, but more restrictive policies on top of that would have been necessary to reverse it. Placer was in between these extremes, with our model suggesting a plateau around October 15th under status quo or the potential for substantial reduction in *R*_*t*_ under business closures. The underprediction bias mentioned earlier is evident here, as Placer did maintain status quo and *R*_*t*_ continued to rise slowly after October 15th. However, the overall trend was still captured; our model forecast was most dire in the areas which turned out to have the worst *R*_*t*_ trend. Policy makers at the time could have used these projections to focus business closures on areas like Fresno, thereby heading off surges in incipient hotspots while reducing economic impact in areas with less worrying trends.

**Figure 3:**
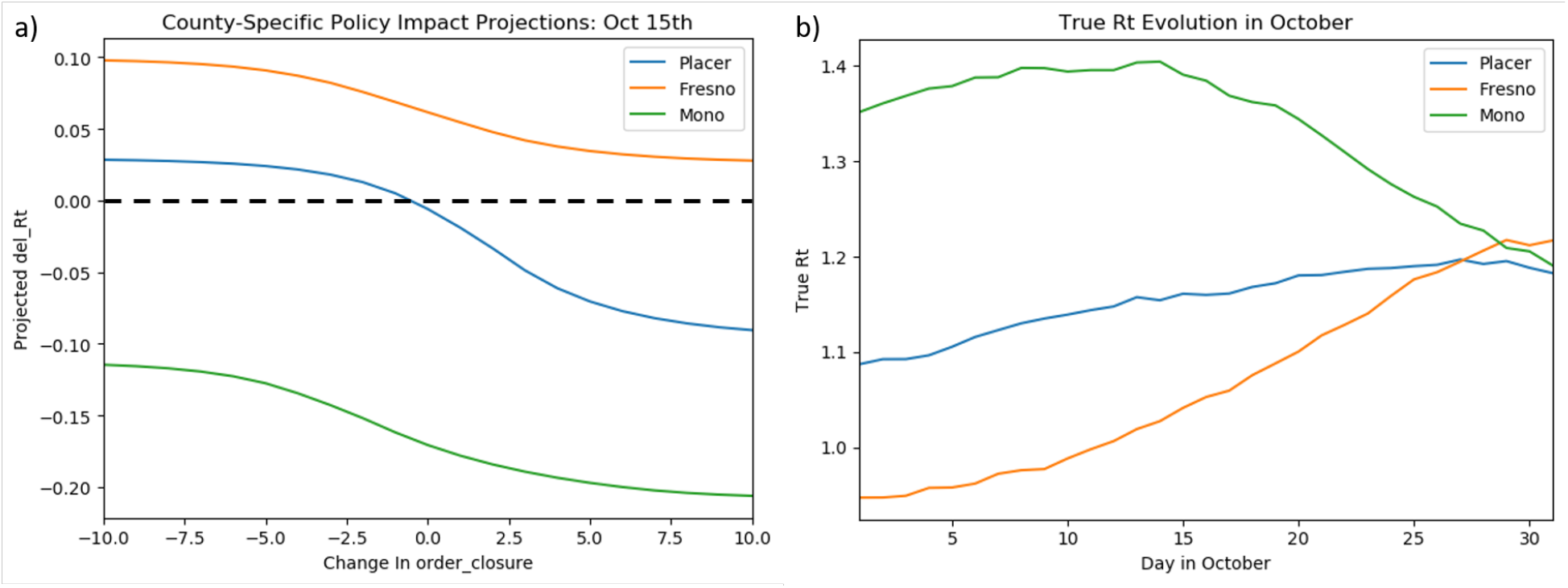
Selected County Projections on October 15th. a) Our model projects that Mono, Placer, and Fresno counties face drastically different policy outlooks on Oct 15th; Mono is on a nice downward trend, Placer is at an inflection point, and Fresno is in the midst of a surge that even maximal business closures couldn’t fully reverse. In reality all three counties maintained loose closure policy for the entire period shown. b) These predictions are well reflected in the true *R*_*t*_ curves from that time.

## Data Availability

All data used are available through links referenced in the manuscript

## Acknowledgments

We thank Dr. Jeremy Goldhaber-Fiebert for providing us with early access to the SC-COSMO public health policy data set, which helped kick-start this research effort.

